# Supervised Machine Learning for the Early Prediction of Acute Respiratory Distress Syndrome (ARDS)

**DOI:** 10.1101/2020.03.19.20038364

**Authors:** Sidney Le, Emily Pellegrini, Abigail Green-Saxena, Charlotte Summers, Jana Hoffman, Jacob Calvert, Ritankar Das

## Abstract

**Purpose:** Acute respiratory distress syndrome (ARDS) is a serious respiratory condition with high mortality and associated morbidity. The objective of this study is to develop and evaluate a novel application of gradient boosted tree models trained on patient health record data for the early prediction of ARDS.

**Materials and Methods:** 9919 patient encounters were retrospectively analyzed from the Medical Information Mart for Intensive Care III (MIMIC-III) data base. XGBoost gradient boosted tree models for early ARDS prediction were created using routinely collected clinical variables and numerical representations of radiology reports as inputs. XGBoost models were iteratively trained and validated using 10-fold cross validation.

**Results:** On a hold-out test set, algorithm classifiers attained area under the receiver operating characteristic curve (AUROC) values of 0.905, 0.827, 0.810, and 0.790 when tested for the prediction of ARDS at 0-, 12-, 24-, and 48-hour windows prior to onset, respectively.

**Conclusion:** Supervised machine learning predictions may help predict patients with ARDS up to 48 hours prior to onset.

## INTRODUCTION

Acute respiratory distress syndrome (ARDS) is a clinical syndrome characterized by hypoxemia in the presence of non-cardiogenic pulmonary edema, and is associated with severe inflammation.^1^ ARDS is estimated to affect at least 190,000 patients per year in the United States^2^ and has been cited as one of the leading causes of admission to the intensive care unit (ICU)^3,4^, with mortality rates ranging between 30%-55%.^5^ The wide variation in reported incidence^6^ and mortality rates^2,7-14^ may relate to difficulties in the recognition and diagnosis of ARDS. Despite high mortality rates and high rates of ICU utilization associated with ARDS, it is still critically misdiagnosed and underdiagnosed in intensive care units on a global scale.^1,5,15^

The broad variability that exists in ARDS diagnosis may be partly explained by differences in risk factors and etiologies, the availability of diagnostics, the quality of chest radiographs and variability of clinician interpretation of radiographs, as well as general clinician ability to recognize ARDS.^7,16^ The inability of healthcare providers to process the volume of clinical data generated while caring for critically ill patients has been cited as another potential reason for poor ARDS recognition.^17,18^ In addition, there is a recognition of the complexity that the clinical and biological heterogeneity within ARDS adds to recognition of the syndrome.^19-27^ Efforts to improve patient outcomes in intensive care settings are often attenuated by a lack of viable methods of early ARDS identification, and have failed to translate to significant reductions in ARDS morbidity and mortality. The most recent Berlin definition^28^ of ARDS was developed in 2012 in response to issues regarding the reliability and validity of the 1994 American-European Consensus Conference (AECC) definition.^29^ Although the Berlin definition has addressed many of the limitations of the AECC definition,^28-30^ defining ARDS in diverse clinical settings remains dependent on some subjectivity of the diagnosing clinician.^31^ It has been suggested that clinicians’ ability to separate ARDS from other heterogeneous causes of respiratory failure is limited,^30,32,33^ and that it can often be difficult to diagnose ARDS in patients who have underlying medical problems with similar symptoms.^34^

Because ARDS treatment options have limited efficacy, there is an interest in identifying patients most at risk of developing ARDS for early prevention strategies, such as antiplatelet therapy,^7,35-37^ as well as early clinical trial enrollment.^38,39^ The opportunity for preventing ARDS onset is constrained to a narrow window.^7,37^ The median time to onset of ARDS is reported to be an average of 2 days after hospital admission.^7,40^ Despite advances in our understanding of ARDS pathogenesis and in the identification of ARDS biomarkers, no single or combination of clinical or biological biomarker has been shown to reliably predict ARDS.^20,41,42^ Therefore, developing clinical decision support (CDS) methods to assist clinicians in the accurate and early prediction of ARDS is a valuable approach to improve patient monitoring, diagnosis, treatment, and outcomes.

CDS technologies have the ability to differentiate between groups of patients with similar conditions, and have been reported to be useful in informing treatment decisions and improving patient outcomes.^41,43^ They have recently been proposed as a method to improve early ARDS detection.^17,44,45^ Through informed data analysis, CDS models can analyze relevant patient data from large electronic health record (EHR) databases and identify cohorts of patients with similar disease progression. Here, we describe the development and analysis of a novel application of supervised machine learning model CDS for the early prediction of ARDS.

## MATERIALS AND METHODS

### Data selection

Data were obtained from the Medical Information Mart for Intensive Care III (MIMIC-III) database, which consists of the inpatient ICU encounters at Beth Israel Deaconess Medical Center between 2001 and 2012.^46^ The MIMIC-III publication states that, “the project was approved by the Institutional Review Boards of Beth Israel Deaconess Medical Center (Boston, MA) and the Massachusetts Institute of Technology (Cambridge, MA). Requirement for individual patient consent was waived because the project did not impact clinical care and all protected health information was deidentified.”^46^ To ensure consistent encoding of data, only data collected with the MetaVision clinical information system were used. All patient data collected using MetaVision was from patients admitted during or after 2008.

We applied additional inclusion criteria (**Figure 1**) to focus the scope of our study. Only patients with age data available and at least 18 years of age were included. Patient stays that did not have at least one observation of each required measurement type (see below) were excluded. Finally, we included only patient stays that had durations within a specified window. The upper limit on length of stay was set at 1000 hours (approximately 41.7 days), in order to account for outliers and transcription errors. The lower limit was dependent on lookahead, and the final study population sizes are listed in **Table 1**. For example, to predict for up to 48 hours before onset of ARDS using a five-hour window, 53 hours of patient data are required for inclusion. Note that, in order to simulate the use case as a screening tool for the general population, we did not require patients to be mechanically ventilated in this study, unlike other similar ARDS studies such as Taoum et al.^47^ and Neto et al.^48^ We have also analyzed separately a subpopulation in which patients are required to have experienced at least one hour of mechanical ventilation to be included in the study population (**Supplemental Table 1**).

**Table 1.** Inclusion table of subjects included in analysis.

| Requirement | Offset |  |  |  |
| --- | --- | --- | --- | --- |
|  | 0 | 12 | 24 | 48 |
| All MIMIC-III encounters | 53432 | 53432 | 53432 | 53432 |
| Age exists, age at least 18 | 53332 | 53332 | 53332 | 53332 |
| Metavision | 24048 | 24048 | 24048 | 24048 |
| At least 1 observation of each required measurement | 23502 | 23502 | 23502 | 23502 |
| Qualifying stay duration | 23328 | 21278 | 16352 | 9919 |

**Figure 1.**
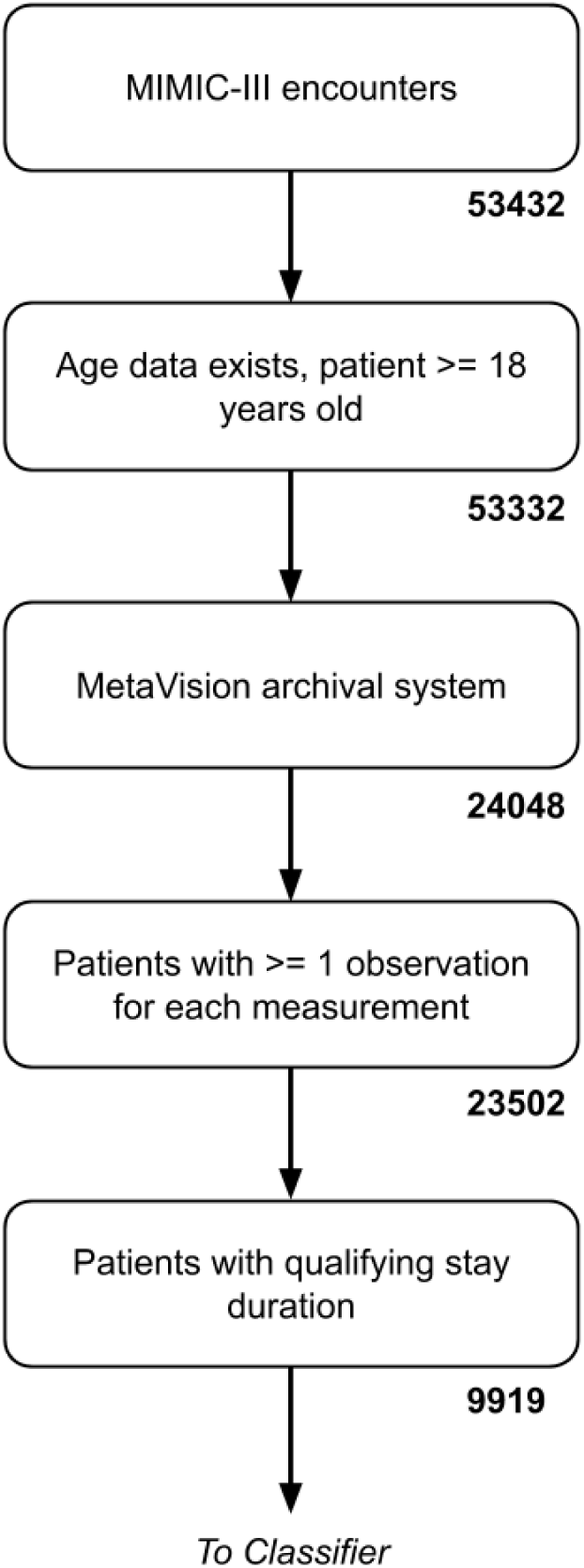
Inclusion criteria for patient encounters in the MIMIC-III dataset. The final inclusion criteria is dependent on prediction lookahead; the value presented here reflects the 48-hour prediction, which filters most stringently.

### Data extraction

Beginning at the first recorded measurement, raw measurements entered into the EHR for each patient stay were binned into one-hour intervals and averaged or summed within bins to produce a single, summarizing value per interval. Antibiotics, urine output, dobutamine, dopamine, epinephrine, norepinephrine, and phenylephrine measurements were summed, and all other clinical measurements listed in **Supplemental Table 2** were averaged. Encoding the data in this way transformed the measurements into discrete time series with consistent time steps, which were more readily handled by the algorithm. Not all raw measurements were available at all hours, so missing values were filled using last-one carry forward imputation. This is a natural imputation method for clinical measurements; observations of a raw measurement are expected to be dependent on the previous observations.^49,50^

**Table 2.**
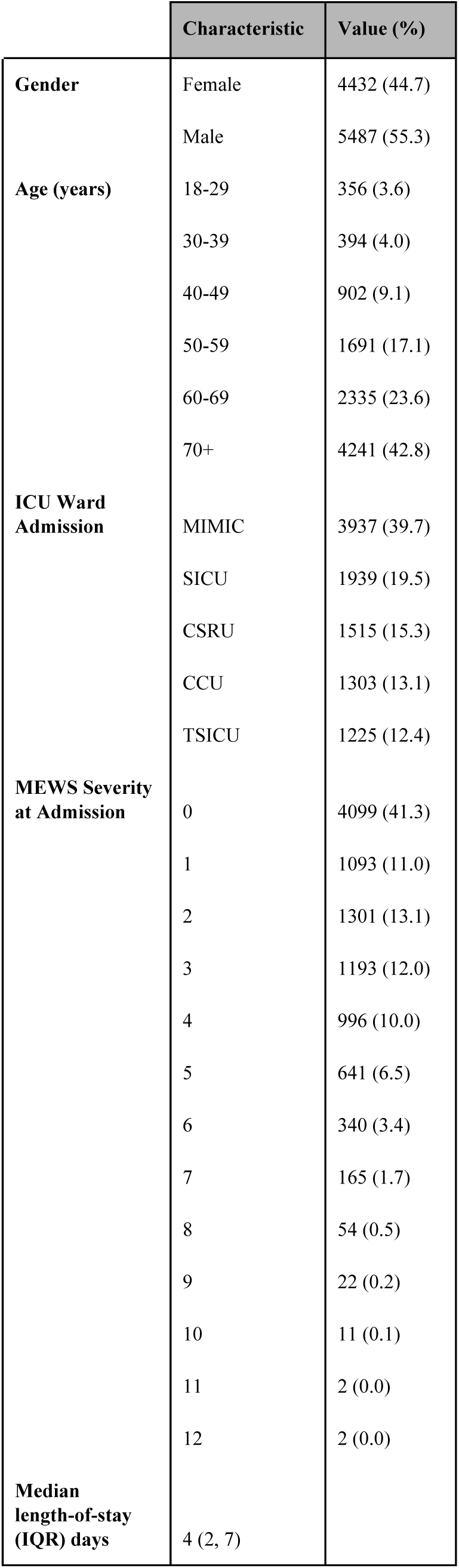
Demographics of subjects included in analyses. Percentage values may not add to 100 due to rounding. Demographics calculated for patients with stay durations of at least 48 hours.

|  | Characteristic | Value (%) |
| --- | --- | --- |
| **Gender** | Female | 4432 (44.7) |
|  | Male | 5487 (55.3) |
| **Age (years)** | 18-29 | 356 (3.6) |
|  | 30-39 | 394 (4.0) |
|  | 40-49 | 902 (9.1) |
|  | 50-59 | 1691 (17.1) |
|  | 60-69 | 2335 (23.6) |
|  | 70+ | 4241 (42.8) |
| **ICU Ward Admission** | MIMIC | 3937 (39.7) |
|  | SICU | 1939 (19.5) |
|  | CSRU | 1515 (15.3) |
|  | CCU | 1303 (13.1) |
|  | TSICU | 1225 (12.4) |
| **MEWS Severity at Admission** | 0 | 4099 (41.3) |
|  | 1 | 1093 (11.0) |
|  | 2 | 1301 (13.1) |
|  | 3 | 1193 (12.0) |
|  | 4 | 996 (10.0) |
|  | 5 | 641 (6.5) |
|  | 6 | 340 (3.4) |
|  | 7 | 165 (1.7) |
|  | 8 | 54 (0.5) |
|  | 9 | 22 (0.2) |
|  | 10 | 11 (0.1) |
|  | 11 | 2 (0.0) |
|  | 12 | 2 (0.0) |
| **Median length-of-stay (IQR) days** | 4 (2, 7) |  |

For each patient stay, we took the vector of measurements at prediction time and concatenated the measurements for up to four hours before the prediction time as additional features. For example, the feature vector for a given patient would consist of all of her measurements at prediction time, followed by all of her measurements for the hour prior, and so on up to four hours before prediction time. Where appropriate, we also concatenated the differences in measurement values between those time steps. In this way, at prediction time, a supervised machine learning technique such as gradient boosted tree ensembles is able to access trend information and covariance structure with respect to time windows. This procedure of transforming time series problems into supervised learning problems has been used in our previous work.^51^

Models were developed using quantitative clinical features taken from the patient EHR, but patients were only required to have age, heart rate, respiratory rate, temperature, diastolic and systolic blood pressure, and SpO2 available. Other quantitative clinical features were included if available, and replaced with “missing” values where not available. Quantitative clinical features included for analysis in patient subpopulations with no required mechanical ventilation, and with at least one hour of mechanical ventilation, are listed in **Supplemental Table 2**. The machine learning algorithm which we applied in this study is capable of learning from the distribution of missing values and can still gain information from relatively sparse features. We included only those patient stays which contained at least one measurement of each of the required features.

We extracted radiology reports and preprocessed them for use in our algorithm. Radiology reports are not expected to be present for every patient stay; moreover, it is valuable information if a patient stay does not have any radiology reports generated. Radiology reports contain complex information concerning clinician insight and the health of a patient. If the reports were present, our experimental design was able to access that complex information for machine learning, and if the reports were not present, the MLA was able to learn information about the lack thereof. Using the Doc2Vec text encoding scheme,^52^ radiology reports were converted into numerical feature vectors. The Doc2Vec encoding network uses the relationships between words and their neighbors, as well as the relationship between paragraphs within a text, to generate a numerical embedding. These embeddings are crucial features in our experimental design for similar reasons; they allow the machine learning algorithm to access text representations of the clinical reality. These numerical embeddings are able to retain much of the relational structure of the text as a feature vector, without necessarily having to retain information about the literal text. The Doc2Vec encoding network, as implemented in the Python package gensim,^53^ was trained on tokenized training texts, preprocessed to remove numbers and non-alphanumeric characters. This corpus of training texts was composed of 117,902 radiology reports, drawn from our training data. Once all training texts were observed and network weights updated, training procedures were frozen, and we then used the fully-trained Doc2Vec encoding network to infer the feature vectors for all of the radiology reports, similarly tokenized and preprocessed. These feature vectors were concatenated onto the existing quantitative clinical variables for patient stays where radiology reports were available. For patient stays where radiology reports were not available, vectors of the same size, containing missing values, were concatenated to the existing variables.

### Gold standard and definition of onset time

In order to generate gold standard labels for ARDS, we followed the Berlin definition^28^ as operationalized in Neto et al.^48^ By examining the patient data for the co-occurrence of positive end expiratory pressure (PEEP) above or equal to 5 cmH_2_O and PaO_2_/FiO_2_ ratio (P/F ratio) below or equal to 300 mmHg, we encoded positive class labels as 1 and negative class labels as 0. The time of first co-occurrence is set as the onset time for ARDS, and prediction time is set to some number of hours prior to this onset. Thus a model described as a 24-hour model is a model for predicting ARDS 24 hours prior to onset by this co-occurrence definition. In order for an onset to be considered ARDS positive, we require that it is acute, as defined by occurring within seven days of the initiation of mechanical ventilation, and we require the presence of bilateral opacities or infiltrates in the radiology report for that patient. Note that the measurements used to determine this gold standard were not used in development or training of the machine learning algorithm used in this study. In pilot experiments, we were able to verify the implementation of ARDS used in this study reproduced ARDS incidence rates observed in Neto et al.^48^

### Experimental methods

For the purposes of evaluation, we reserved 10% of the patient stays within the MIMIC-III dataset, chosen at random, as a hold-out dataset and used only the remaining 90% to train, validate, and iterate our predictive models. This hold-out data represents unseen new data and can be used to gauge performance of machine learning algorithms in the setting of novel data prediction. Although we are primarily interested in prediction at 24-hours prior to onset, we also trained models for detection of ARDS at onset and prediction of ARDS at 12-hours and 48-hours prior to onset.

All predictive models described in this paper are instances of the XGBoost gradient boosted tree model,^54^ implemented using the Python package. XGBoost is a state-of-the-art tree ensemble method that builds progressively on the loss generated by weak decision tree base learners. XGBoost is capable of learning quickly and effectively from large amounts of data, and is flexible to the point that it is able to learn even from missing data. By making use of this capability, we are able to construct predictive models that do not require radiographs or radiology reports to make meaningful predictions. It is important to note that decision tree models, including tree ensembles, do not make distributional assumptions, and so are well-suited for settings where specifying a generative distribution is difficult.

Three of the available hyperparameters for XGBoost were selected using exhaustive grid search five-fold cross validation, performed exclusively on the training data. Five folds for hyperparameter tuning is the default for hyperparameter grid search due to considerations of computational constraints, as implemented in Scikit-learn.^55^ The hyperparameters tuned are number of base learners, the learning rate, and the maximum depth of a base learner. The hyperparameters were tuned across ranges of values centered around 1000, 0.1, and 5 for number of base learners, learning rate, and maximum depth, respectively. The values selected as the centers were determined by iteratively narrowing the grid search range. These three hyperparameters affect the values the internal model parameters take over the course of training, and thus also significantly contribute to the final model parameters.

The XGBoost predictive models were all iteratively trained and tested using ten-fold cross validation with early stopping mechanisms in order to prevent overfitting. In this validation paradigm, the data was partitioned into ten random segments, or folds. Training occurred on nine of the folds, and the remaining fold was used to monitor performance for overfitting. Each of the ten models trained were then tested on the hold-out test set partitioned prior to hyperparameter tuning, and the final metrics reported were averages for the metrics across the ten models. Metrics reported include area under the receiving operator curve (AUROC), standard deviation for the area metrics, sensitivity, specificity, accuracy, recall, diagnostic odds ratio (DOR), and positive and negative likelihood ratios (LR+ and LR-, respectively). Once all models were trained, we evaluated the performance of the models in predicting the ARDS labels of the hold-out set, and the same performance metrics were reported.

In addition to this main set of experiments validating the effectiveness of our algorithm as a screening tool developed and tested on the general patient population, we conducted an additional analysis in which we developed and evaluated the same algorithm using only mechanically ventilated patients. All procedures, from partitioning into training and hold-out test sets to hyperparameter tuning and training, were performed identically in this additional experiment.

## RESULTS

Using the training data, we performed five-fold cross validation on every combination of hyperparameter values in our pre-specified hyperparameter ranges. In total there were 72 different hyperparameter combinations, and with five-fold cross validation, a total of 360 models were fit on the training data. The evaluation metric used to determine the best performing hyperparameter combination was AUROC. The hyperparameters selected to train the final models were: 1000 base learners, a learning rate of 0.03, and a base learner maximum depth of six partition levels.

Patient data were drawn from the MIMIC III data set, which consists of the inpatient ICU encounters at Beth Israel Deaconess Medical Center and minimally filtered by age, clinical information system, and a small set of required measurements. Demographic data of all patient encounters from the MIMIC-III dataset are presented in **Table 2**, including the distribution of admissions to various wards in the ICU and the distribution of physiologic derangement, represented by MEWS scores, at admission.

ARDS onset detection and prediction performance is summarized by the Receiver Operating Characteristic (ROC) curves in **Figure 2**. ROC curves show sensitivity (the fraction of ARDS positive cases that received an ARDS positive label) as a function of 1-specificity (the fraction of ARDS negative cases that received an ARDS positive label). Operating points of approximately 0.80 sensitivity were selected for each model to facilitate comparisons of performance. Each ROC curve represents the average performance under 10-fold cross validation. The classifier demonstrated an AUROC of 0.905, 0.827, 0.810, and 0.790 for early ARDS prediction on the test set at 0-, 12-, 24-, and 48-hours prior to onset, respectively (**Figure 2**). AUROC curves demonstrated high sensitivity and specificity of algorithm predictions for ARDS onset up to 48 hours in advance on the test set.

**Figure 2.**
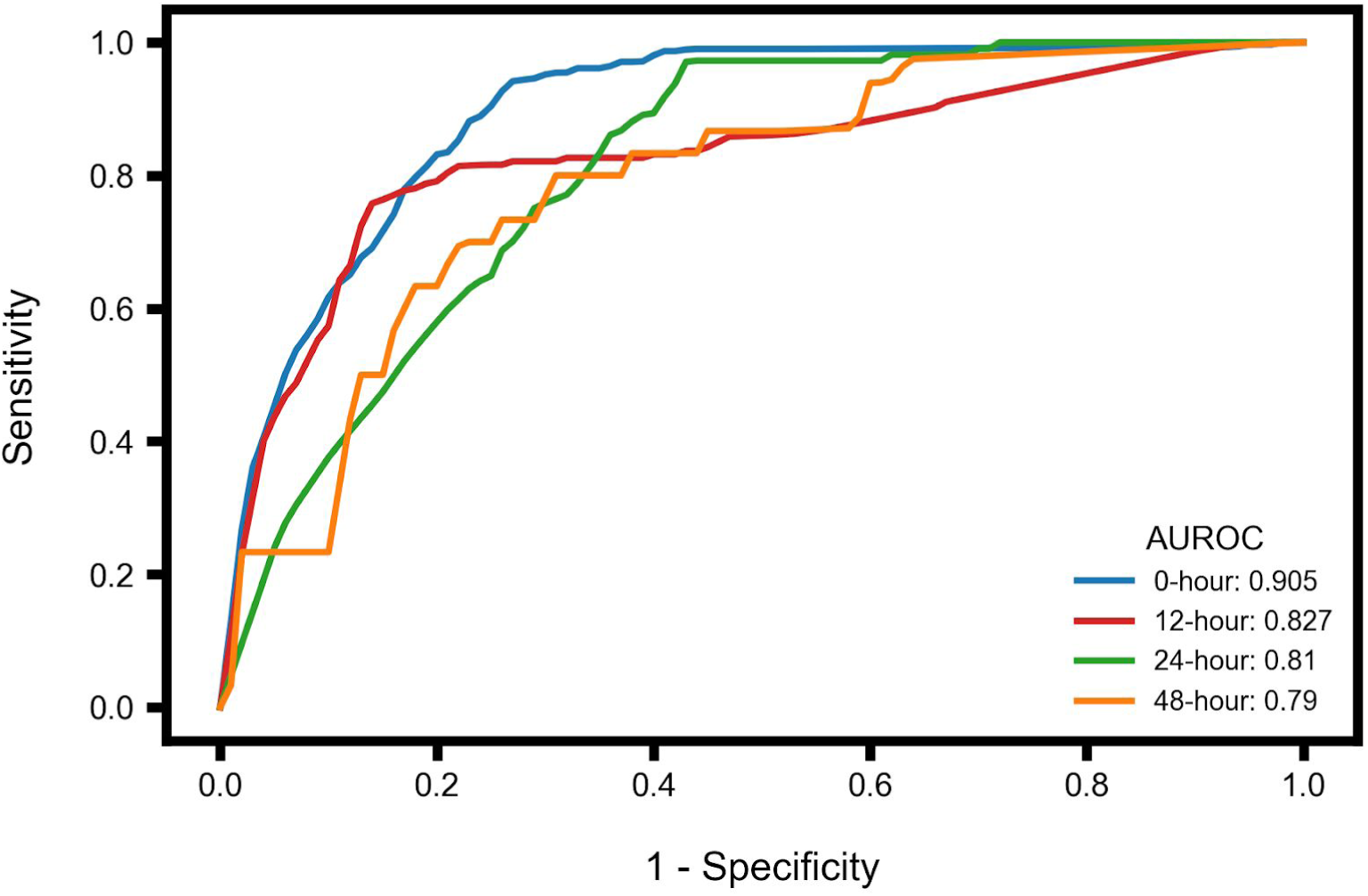
Area Under Receiver Operating Characteristic (AUROC) curves and values for ARDS onset detection and prediction at 12, 24, and 48 hours prior to onset. AUROC performance of XGBoost models on a separate hold-out test set for early ARDS prediction, up to 48 hours prior to onset. Curves are averaged across 10 folds.

Multiple performance metrics are shown in **Table 3**, including AUROC,, sensitivity and specificity, representing a variety of clinically relevant assessments for the general patient population. All metrics were calculated with common operating points near sensitivity = 0.80 in order to allow for direct comparisons. Testing performance metrics for the patient population with at least one hour of mechanical ventilation are reported in **Supplemental Table 3**, and the AUROC curves associated with the performance are shown in **Supplemental Figure 1**. In this mechanically ventilated population, our classifier demonstrated AUROC performance of 0.843, 0.858, 0.810, and 0.790 for early ARDS prediction on the test set at 0-, 12-, 24-, and 48-hours prior to onset, respectively.

**Table 3.** Model performance metrics on the training and testing (hold-out) sets at 0-, 12-, 24-, and 48-hour prediction windows. AUROC: area under the receiving operator curve; DOR: diagnostic odds ratio; LR+ and LR-: positive and negative likelihood ratios, respectively. Values presented are means and standard deviations for the metrics across 10 folds.

|  | Onset | 12-hour | 24-hour | 48-hour |
| --- | --- | --- | --- | --- |
| **AUROC** | 0.905<br>(0.009) | 0.827<br>(0.015) | 0.810<br>(0.035) | 0.790<br>(0.079) |
| **Sensitivity** | 0.806<br>(0.000) | 0.789<br>(0.000) | 0.818<br>(0.000) | 0.667<br>(0.000) |
| **Specificity** | 0.823<br>(0.014) | 0.828<br>(0.052) | 0.683<br>(0.073) | 0.852<br>(0.063) |
| **F1** | 0.109<br>(0.006) | 0.079<br>(0.015) | 0.020<br>(0.018) | 0.015<br>(0.004) |
| **DOR** | 19.477<br>(1.829) | 19.704<br>(5.953) | 10.452<br>(3.664) | 13.175<br>(4.485) |
| **LR+** | 4.576<br>(0.354) | 4.938<br>(1.253) | 2.719<br>(0.666) | 5.058<br>(1.495) |
| **LR-** | 0.235<br>(0.004) | 0.255<br>(0.017) | 0.269<br>(0.028) | 0.393<br>(0.032) |
| **Accuracy** | 0.825<br>(0.010) | 0.839<br>(0.045) | 0.817<br>(0.160) | 0.851<br>(0.061) |
| **Recall** | 0.774<br>(0.000) | 0.732<br>(0.017) | 0.427<br>(0.369) | 0.333<br>(0.000) |

As shown in **Supplemental Table 4**, antibiotics administration appears to yield a significant amount of information about the classifier across all prediction times in the general patient population. However, there are few other observable trends in feature importances that are consistent. It should be noted that the stochastic nature of the XGBoost algorithm, which extends to the subset of columns which it considers in individual trees in its ensemble, limits the interpretability of feature importances.

## DISCUSSION

We have described a method for the early prediction of ARDS using supervised machine learning models. We developed these models using quantitative clinical features extracted from the patient EHR data, including age, heart rate, respiratory rate, temperature, diastolic and systolic blood pressure, SpO2, GCS, antibiotics, creatinine, bilirubin, fluids administration, INR, lactate, pH, platelets, WBC, PP, MAP, and derived organ dysfunction, as well as numerical representations of radiology reports. Our approach circumvents issues associated with keyword-based text analysis by using higher-level representations of the text in radiology reports. These representations are used as features in our model, alongside the patient quantitative, structured data. The use of this structured data to complement radiographic reports softens the limitations on timeliness which may be presented by requiring the use of information derived from chest radiographs. In these ways, the method we describe diversifies and improves upon existing approaches for the prediction of ARDS.

Inability to anticipate which patients are likely to develop ARDS is a major obstacle to early intervention or prevention studies.^56^ Epidemiologic data suggest that the syndrome is rarely present at the time of hospital admission or initial emergency department (ED) evaluation, but develops over a period of hours to days in subsets of at-risk patients.^57-61^ Therefore, evaluating model performance at >24 hours preceding onset is valuable because it facilitates identification of patients who would benefit from ARDS progression and prevention interventions. Alerting systems for the long horizon prediction of ARDS have been validated in similar studies of mechanically ventilated patients and those with moderate hypoxia.^62,63^

Rule-based systems such as that developed by Herasevich et al.^64,65^ have been used to screen patients for ARDS by analyzing patient EHR data.^66^ For example, Lung Injury Prediction Score (LIPS) is a rules-based system used for predicting development of ARDS and mortality.^56,67^ The model allows clinicians to incorporate a series of risk factors to predict patients who will develop ARDS using clinical data at the time of presentation to the emergency department (ED), and it has been shown in a recent study to be a viable method for predicting mortality and ARDS onset in critically ill surgical patients.^30,68^ However, it is unclear how LIPS performs in patients who develop predisposing conditions after initial presentation to the ED.^68^ Interpretation of the LIPS score has also been shown to be highly dependent on the diagnosing clinician, and overall demonstrates suboptimal sensitivity and specificity.^30^ In response to these limitations, alternative systems have been proposed to promote early ARDS recognition and improve patient outcomes.

Non rules-based CDS systems provide a viable alternative for ARDS detection and prediction, as they are capable of efficiently incorporating complex patient data sets and are therefore less reliant on clinician subjectivity. Several studies have focused on the development of ARDS detection systems.^17,19^ Zaglam et al. developed a computer-aided diagnosis system for the assessment of ARDS from chest radiographs^19^, with results demonstrating a sensitivity of 90.6% and a specificity of 86.5%. Reamaroon et al.^17^ used support vector machines (SVM) to account for clinical diagnostic uncertainty when training an algorithm to detect patients who develop ARDS. Results indicated algorithm ability to achieve improvement in detecting patients with ARDS, with an increase of 10% in AUROC when compared with a conventional SVM algorithm.^17^ A retrospective study by Schenck et al. used a machine learning technique to predict for a subphenotype of ARDS in patients at the point of their enrollment into clinical trials.^69^ Other algorithms and text processing tools have also been evaluated for their ability to assist with patient screening for ARDS.^62,70,71^ These studies represent a promising means for addressing the issues clinicians face when identifying pre-existing ARDS. However, there is a pressing need for CDS systems that can predict ARDS onset sufficiently in advance to provide clinicians with time to undertake preventative measures. For example, Taoum et al. described a novel approach for early prediction of ARDS using continuous physiological signals of heart rate, respiratory rate, peripheral arterial oxygen saturation and mean airway blood pressure.^47^ Results indicated that ARDS can be detected in the early phases of occurrence with a sensitivity of 65% and a specificity of 100%, on average 39 hours prior to onset.^47^ While this study was undertaken on a small dataset, which limits generalizability, it demonstrates the ability of machine learning algorithms to predict the onset of ARDS.

The method of Taoum et al. relies on minute-by-minute samples from physiological monitors to detect ARDS, which introduces a potential barrier to hospital integration and which ignores the benefits of unstructured clinical notation data.^47^ In contrast, our method uses relatively sparsely sampled structured data, such as vital signs and lab tests, in addition to unstructured notation data. The method of Zaglam et al. requires chest radiographs to be obtained before it may assess the presence or absence of ARDS, which may hinder the early diagnosis or prediction of ARDS, and does not use text data.^19^ While the rules-based method of Herasevich et al. uses unstructured radiographic report data, it does so by searching reports for a list of keywords, which is vulnerable to misdiagnosis arising from the negation of keywords and potentially neglects more complicated textual indications of ARDS.^64^ In contrast, our use of Doc2Vec enables the extraction of rich, contextual information from unstructured texts, including information concerning chest radiographs highly relevant to ARDS. Our approach does so without explicitly requiring radiographs for inclusion in screening, which allows the tool to be used as a screening tool for the general population.

Our supervised machine learning models demonstrate high diagnostic metrics for ARDS recognition and prediction (**Table 3**). The testing curves of **Figure 2** demonstrate the model’s strength in diagnosis at the time of ARDS onset, with an AUROC value 0.905 for the general patient population. These metrics outperform those reported in other studies.^69^ While the quality of diagnostic metrics decay as they are made increasingly early prior to ARDS onset, the 12-hour prediction of ARDS offers operating points with high sensitivity and specificity. **Table 3** illustrates a clinically relevant operating point with sensitivity of 0.806 and specificity of 0.823. Increasing the recognition of ARDS onset in clinical settings is essential for improving the management of patients with ARDS, and enabling clinical trial enrollment to improve ARDS treatment methods. Early prediction of ARDS onset offers opportunities for increased patient monitoring, possible prevention,^35-37^ and the development of novel preventative measures.

In the mechanically ventilated subpopulation, our supervised machine learning models demonstrated a similarly high level of diagnostic performance for ARDS recognition and prediction (**Supplemental Table 3**). Models in both the mechanically ventilated subpopulation and the general population achieved high sensitivity and specificity for 12-hour prediction of ARDS, with an operating point with sensitivity of 0.778 and specificity of 0.810 in the mechanically ventilated subpopulation. At time of ARDS onset in this subpopulation, an AUROC of 0.843 was observed in this subpopulation compared to AUROC of 0.905 in the general population. The performance 12 hours prior to onset was higher in the mechanically ventilated subpopulation, with an AUROC of 0.858 compared to AUROC of 0.827. Overall we observed similar performance in both patient populations.

There are several limitations to the study. The results of the study may not directly apply to a different definition or operationalization of ARDS, as we used the Berlin definition for labeling patients as ARDS-positive and ARDS-negative. This study was a retrospective analysis, which may not translate to prospective improvements in clinical settings. In particular, the performance metrics we report do not capture the interaction of clinicians with the information such a tool would provide, or the limitations of ARDS prevention and treatment options. Finally, this study concerned a single-center study of ICU data and therefore the results may not translate to other clinical settings or wards, especially wards of less intensive care. In future work we hope to develop and evaluate this tool in a variety of live clinical settings.

## CONCLUSION

This analysis demonstrates the use of a gradient boosted tree model for the early prediction and identification of ARDS using retrospective patient data. The algorithm developed in this study may assist both in recruitment for ARDS clinical trials and the improved prediction and early recognition of ARDS.

## Data Availability

Data are publicly available.

## Author contributions

RD had full access to all of the data in the study and takes responsibility for the integrity of the data and the accuracy of the data analysis. Conceptualization, SL, JC, JH, CS, RD; Methodology, SL, JC, RD; Formal Analysis, SL, JC; Investigation, SL, JC; Writing--Original Draft, RD, JH, JC, SL, AGS, CS, and EP; Supervision, RD and JH.

## Abbreviations

AECC: American-European Consensus Conference
APACHE: Acute Physiologic Assessment and Chronic Health Evaluation
ARDS: Acute Respiratory Distress Syndrome
AUROC: Area Under Receiver Operating Characteristic
CDS: Clinical Decision Support
DOR: Diagnostic Odds Ratio
ED: Emergency Department
EHR: Electronic Health Records
GCS: Glasgow Coma Scale
ICU: Intensive Care Unit
ICD-9: International Statistical Classification of Diseases version 9
INR: International Normalised Ratio
LIPS: Lung Injury Prediction Score
LR+: Positive Likelihood Ratio
LR-: Negative Likelihood Ratio
MAP: Mean Arterial Pressure
MIMIC III: Medical Information Mart for Intensive Care version III
MLA: Machine Learning Algorithm
PEEP: Positive End Expiratory Pressure
P/F ratio: PaO2/FiO2 ratio
PP: Pulse Pressure
ROC: Receiver Operating Characteristic
SAPS: Simplified Acute Physiology Score
SOFA: Sequential Organ Failure Assessment
WBC: White Blood Cell Count

